# Self-reported safety of the BBIBP-CorV (Sinopharm) COVID-19 vaccine among Iranian people with multiple sclerosis

**DOI:** 10.1101/2021.10.17.21265114

**Authors:** Masoud Etemadifar, Amir Parsa Abhari, Hosein Nouri, Amirhosein Akhavan Sigari, Seyed Mohammad Piran Daliyeh, Mohammad Reza Maracy, Mehri Salari, Shiva Maleki, Nahad Sedaghat

## Abstract

To affirm the short-term safety of the BBIBP-CorV (Sinopharm) COVID-19 vaccine among people with multiple sclerosis (pwMS), 517 vaccinated and 174 unvaccinated pwMS were interviewed. 16.2% of the vaccinated pwMS reported at least one neurological symptom in their respective at-risk periods (ARP) – a period from the first until two weeks after the second vaccine dose. In a multivariable logistic regression model, presence of comorbidities (P = 0.01), being on natalizumab (P = 0.03), and experiencing post-vaccination myalgia (P < 0.01) predicted the development of post-vaccination neurological symptoms. One MS relapse, one COVID-19 contraction, and one ulcerative colitis flare after the first, and four MS relapses after the second dose, were the only reported serious adverse events during the ARPs. A multivariable Poisson regression model accounting for possible confounders failed to show any statistically-significant increase in relapse rates during the ARPs of vaccinated, compared to the prior year of unvaccinated pwMS (P = 0.78). Hence, the BBIBP-CorV vaccine does not seem to affect short-term MS activity. Furthermore, as 83.33% of the unvaccinated pwMS reported fear of possible adverse events to be the reason of their vaccination hesitancy, provision of misinformed pwMS with evidence-based consultations in this regard is encouraged.

## Introduction

Nearly a year passed the emergence of the coronavirus disease 2019 (COVID-19), several vaccines were developed and approved. The BNT162b2 (Pfizer-Biontech), mRNA-1273 (Moderna), ChAdOx1 (AstraZeneca), and BBIBP-CorV (Sinopharm) vaccines are currently being used worldwide and all have received authorization from the world health organization (Coronavirus Disease (COVID-19)).

Vaccination is considered as the most prominent scheme to halt the rapid surge of COVID-19 and end the pandemic; However, administration of vaccines in people with multiple sclerosis (pwMS) has always been a matter of controversy. These concerns arose considering the potential effect of disease-modifying therapies (DMTs) on the immunogenicity of the vaccines or probable vaccine-associated worsening of the disease course. Theoretically, vaccines may either trigger and increase the risk of MS in healthy individuals, or increase relapses in already-diagnosed pwMS, as seen in the controversial cases of yellow fever vaccines (Farez and Correale 2011; Reyes et al. 2020). Nevertheless, no other population-level association has been identified between vaccination and MS development or relapses (Confavreux et al. 2001). Moreover, this seems to be also true for COVID-19 vaccines based on the limited data available to date (Achiron A., Dolev M., et al. 2021; Ali Sahraian et al. 2021). Based on our observations in the clinic however, pwMS are concerned about the safety of COVID-19 vaccination, considering their disease. Therefore, providing evidence-based consultations and reasoning has gained more importance among them.

Inadequate data was available at the time of initiation of this study on the safety of the BBIVP-CorV COVID-19 vaccine in pwMS. Therefore, this preliminary study aimed to assess the BBIVP-CorV vaccine’s short-term safety in a relatively large cohort of pwMS in Iran.

## Materials and Methods

Following the STROBE guidelines, a questionnaire, first created by a team of researchers, was used to collect information from the pwMS. These patients were identified and assessed for eligibility via an online platform (Porsline.ir), distributed in vaccination centers and MS forums across Iran from June 7^th^ until 12^th^, 2021, in which they entered their information, including their telephone number. A trained nurse collected the information on eligible patients via telephone calls, from June 18^th^ until 24^th^, 2021. Patients who did not provide their telephone numbers were excluded. Eligible participants were pwMS with a minimum disease duration of one year who either received the BBIVP-CorV vaccine or no vaccine. Prior sample size calculation was not performed, and therefore, no goal was set for the sample size.

In the vaccinated cohort, outcomes were obtained pertaining to an at-risk period (ARP) from the first dose until 2 weeks after the second, and in the unvaccinated cohort, pertaining to a one-year period before the study. All variables and their measurement are summed up in Table 1. An MS relapse was defined as development of new, or worsening of pre-existing neurological symptoms, lasting for at least 24 hours without concurrent fever/infection, preceded by 30 days of neurological improvement/stability.

**Table 1;.** Study variables and their measurement

| Variables | Measurement |
| --- | --- |
| Primary outcome |  |
| 1. Number of MS relapses | Count |
| Secondary outcomes |  |
| 2. General post-vac symptoms | Categorical |
| 3. Neurological post-vac symptoms | Categorical |
| 4. Possible serious events | Count |
| 5. Reason for vac-hesitancy | Nominal |
| Other |  |
| 6. Duration since the first dose | Number of weeks |
| 7. Age | Years |
| 8. Sex | Female/male |
| 9. Disease duration | Years |
| 10. MS type | Relapsing-remitting/progressive |
| 11. Disease-modifying therapy | Categorical |
| 12. Number of Comorbidities | Count |
| 13. Previous COVID-19 | Binary |
| Abbreviations: vac, vaccination; MS, multiple sclerosis; |  |

Baseline characteristics of the vaccinated and unvaccinated cohorts were compared using proper (non)parametric statistical tests. Among the vaccinated pwMS, a multivariable logistic regression model was used to investigate the effect of age, sex, presence of comorbidities, MS type and duration, DMTs, and presence each general post-vaccination symptoms on development of neurological post-vaccination symptoms. The measures pertaining to the primary outcome i.e. MS relapse rate, were compared between the vaccinated and unvaccinated cohorts using a multivariable Poission regression model accounting for age, sex, MS duration, MS type, DMT, and offset for observation period i.e. the ARP for the vaccinated and the prior year for the unvaccinated pwMS. A two-tailed P value of 0.05 was considered as the threshold for diagnosis of statistical significance. Analyses were carried out using SPSS23 (IBM Inc.) for MacOS.

## Results

Figure 1 summarizes the participant flow of the study. Table 2 summarizes the general baseline characteristics, and Table 3 the post-vaccination symptoms of the participants. The vaccinated and unvaccinated cohorts differed regarding their duration of MS (P < 0.01), previous COVID-19 history (P < 0.01) and their DMTs (P = 0.05). A greater portion of pwMS were on no DMTs in the unvaccinated cohort, and a greater portion were on high-efficacy DMTs in the vaccinated cohort.

**Table 2;.** General characteristics of participants

| General Characteristics | Vaccinated (n = 517) | Unvaccinated (n = 174) | P |
| --- | --- | --- | --- |
| Mean age (SD) | 37.81 (8.74) | 36.75 (10.20) | 0.19 |
| Sex (female/male) | 397 / 120 | 137 / 36 | 0.18 |
| Presence of comorbidities (Yes/No) | 101 / 416 | 31 / 143 | 0.62 |
| Median MS duration (Range) | 8 (29) | 6 (20) | <0.01 |
| MS phenotype (R/P) | 427 / 90 | 154 / 20 | <0.01 |
| History of COVID-19 (Yes/No) | 208 / 309 | 28 / 146 | 0.07 |
| DMTs (n, %) |  |  | 0.05 |
| • No DMT | 20 (3.87) | 14 (8.04) |  |
| • Interferons | 161 (31.14) | 50 (28.74) |  |
| • Glatiramer Acetate | 53 (10.25) | 22 (12.64) |  |
| • Dimethyl Fumarate | 100 (19.34) | 41 (23.56) |  |
| • Teriflunomide | 41 (7.93) | 17 (9.77) |  |
| • Fingolimod | 48 (9.28) | 14 (8.05) |  |
| • Natalizumab | 4 (0.77) | 1 (0.57) |  |
| • Rituximab | 90 (17.41) | 15 (8.62) |  |
| Vaccination status |  | NA |  |
| • One dose | 100 (19.3) |  |  |
| • Two doses | 417 (80.7) |  |  |
| Median ARP (Range) | 6 (4) |  |  |
| Abbreviations: SD, standard deviation; R, relapsing; P, progressive; DMT, disease-modifying therapy; ARP, at-risk period; NA, not applicable. |  |  |  |

**Table 3;.**
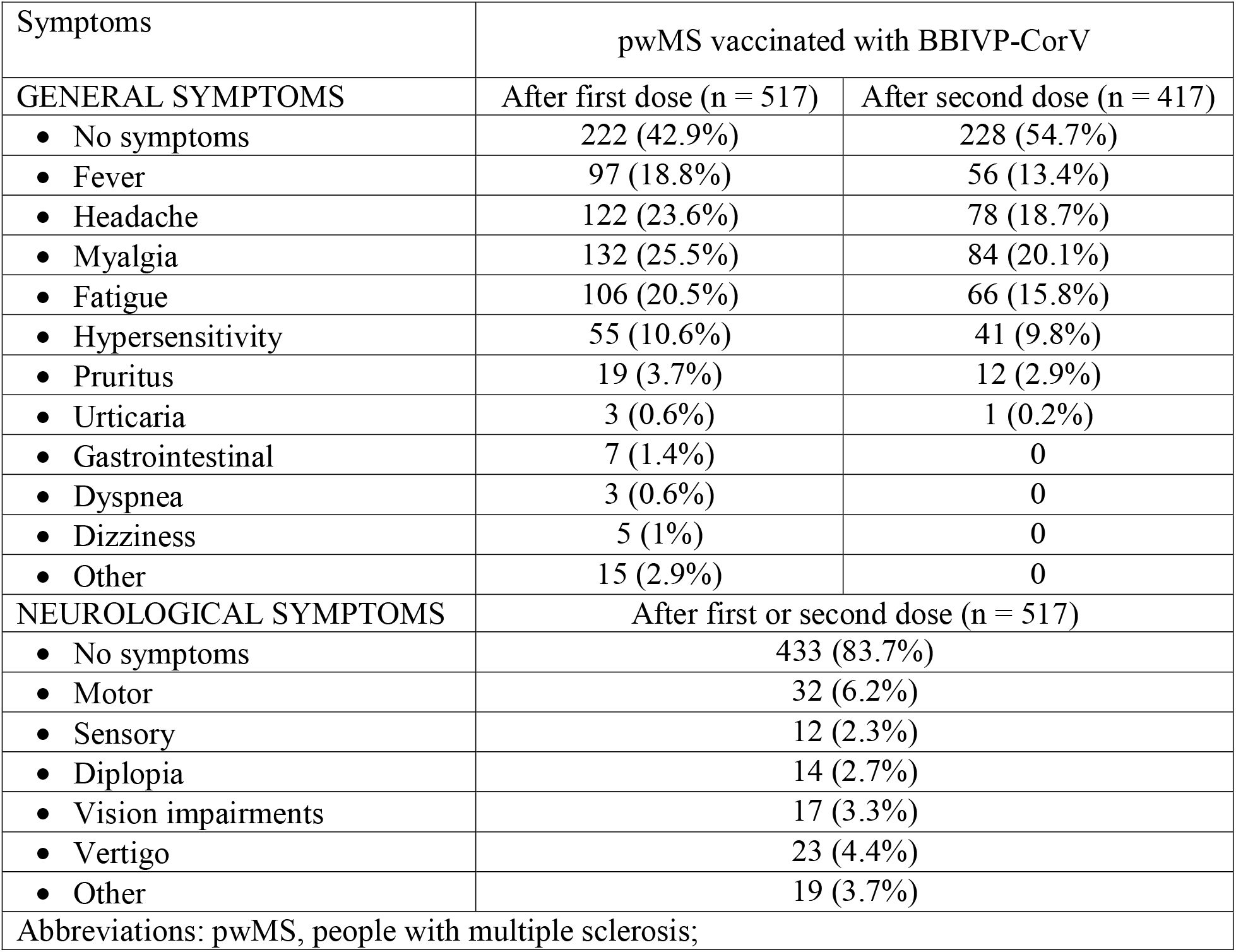
Post-vaccination symptoms of participants during the ARPs.

**Fig. 1;.**
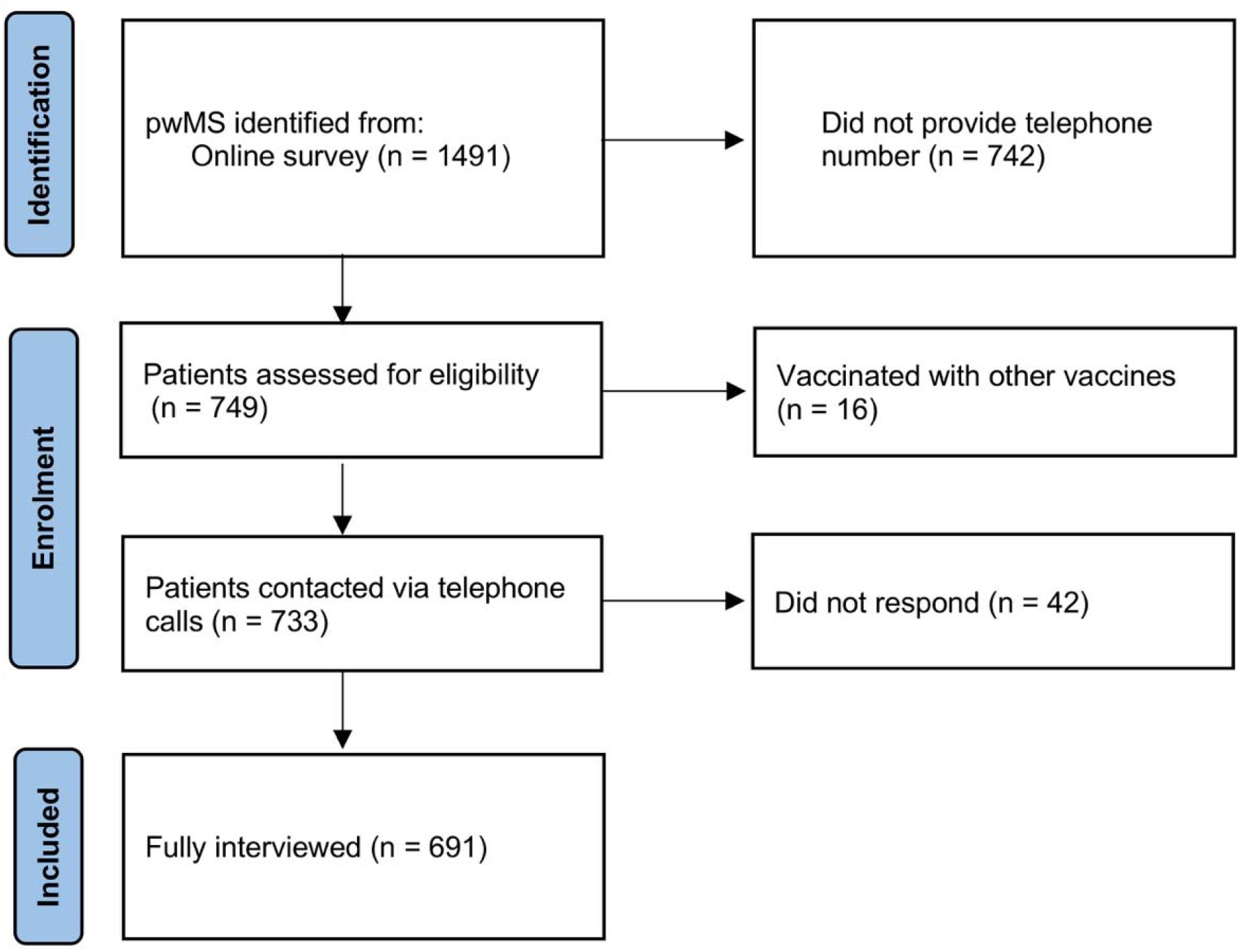
Study flow diagram.

84 (16.2%) of the vaccinated participants experienced at least one post-vaccination neurological symptom, among which motor symptoms and vertigo were more common (Table 3). Presence of comorbidities (B [SE]: 0.76 [0.30], P = 0.01), being on natalizumab therapy (B [SE]: 2.63 [1.24], P = 0.03), and experiencing post-vaccination myalgia (B [SE]: 0.73 [0.28], P < 0.01), predicted the development of post-vaccination neurological symptoms in the multivariable logistic regression model. Only five vaccinated women with MS (two secondary progressive and three relapsing-remitting) experienced neurological symptoms during the ARP that met the criteria for an MS relapse, two were on dimethyl fumarate, one on glatiramer acetate, one on fingolimod, and one on rituximab therapy. One of the relapses followed the first and four followed the second dose of the vaccine. All relapses were followed by either partial or complete improvement after treatment with corticosteroids. The multivariable Poisson regression model did not indicate any significant difference between the relapse rates of vaccinated pwMS in the ARP and the unvaccinated pwMS in the prior year (Table 4). Furthermore, apart from one case of COVID-19 and one case of ulcerative colitis flare – both after the first dose – no other serious adverse events were reported.

**Table 4;.** Results of multivariable Poisson regression.

| Predictors | Multivariable Poisson regression (n = 691, outcome: MS relapse) |  |  |
| --- | --- | --- | --- |
|  | B | SE | P |
| Age (per year) | -0.037 | 0.026 | 0.157 |
| Sex |  |  |  |
| - Male | (ref) |  |  |
| - Female | -0.094 | 0.576 | 0.871 |
| History of COVID-19 |  |  |  |
| - No | (ref) |  |  |
| - Yes | 0.404 | 0.564 | 0.473 |
| MS type |  |  |  |
| - Relapsing | (ref) |  |  |
| - Progressive | 0.160 | 0.723 | 0.824 |
| MS duration (per year) | 0.054 | 0.050 | 0.283 |
| DMT |  |  |  |
| - No DMT | (ref) |  |  |
| - Injectables (IFN, GA) | -0.251 | 1.200 | 0.834 |
| - Orals (DMF, TFN, FNG) | 0.565 | 1.116 | 0.612 |
| - Infused (RTX, NTZ) | 0.541 | 1.214 | 0.656 |
| Cohort |  |  |  |
| - Unvaccinated | (ref) |  |  |
| - Vaccinated | 0.163 | 0.579 | 0.779 |
| Abbreviations: MS, multiple sclerosis; ref, reference; DMT, disease-modifying therapy; IFN, interferons; GA, glatiramer acetate; DMF, dimethyl fumarate; TFN, teriflunomide; FNG, fingolimod; RTX, rituximab; NTZ, natalizumab. |  |  |  |

Among the unvaccinated pwMS, the main reason for vaccination hesitancy was reported to be fear of possible adverse events (83.33%). Other reasons included pregnancy and lactation-related reasons (4.60%), believing that they have already obtained immunity after contracting COVID-19 (4.02%), waiting for other types of vaccines (1.72%), fears of conspiracy (2.30%), disbelieving vaccination (3.45%), and unawareness of eligibility (0.57%).

## Discussion

Our results suggest that the BBIVP-CorV vaccine does not increase the risk of relapse in pwMS, although a relatively small proportion of vaccinated pwMS may experience neurological symptoms. 83.7% of our participants did not develop any neurological sequelae following vaccination, and only 6 (1.2%) experienced post-vaccination MS relapses. However, we acknowledge the probability of possibly missed relapses and the short observation period of our study, which could have accounted for the relatively low relapse frequency in our study. In a relatively similar cohort of 555 pwMS vaccinated with BNT162b2, followed up 14-21 days after the second dose of the vaccine, 13 (2.3%) acute MS relapses were reported (Achiron A., Dolev M., et al. 2021). In another study on 29 pwMS vaccinated with ChAdOx1 and four with BNT162b2, no lasting neurological adverse event or MS relapse was observed in at least two months of follow-up (Allen-Philbey et al. 2021). Similar to the present study, a more recent study on Iranian pwMS receiving the first dose of BBIBP-CorV also did not find an association between vaccination and short-term MS activity, although five post-vaccination MS relapses were reported (Ali Sahraian et al. 2021). Whether or not the mentioned relapses could be attributed to COVID-19 vaccines is unclear.

The BBIBP-CorV is an inactivated virus, administered in a two-dose regimen (Wang et al. 2020). Its phase 1 and 2 trials and a study on a cohort of 89 non-MS individuals found no neurological adverse event or serious side effect (Abu-Hammad et al. 2021; Xia et al. 2021). In its phase 3 trial, apart from injection-site reactions, myalgia and headache were the most frequent systemic adverse events; five individuals experienced more severe systemic adverse events affecting the nervous system (Al Kaabi et al. 2021). Systemic adverse events were more frequent after the first dose than the second in our study. Nevertheless, numerous studies have shown promising results in pwMS following the administration of inactivated vaccines (e.g., influenza or tetanus toxoid vaccines) (Kaufman et al. 2014; Olberg et al. 2018; Metze et al. 2019).

The practical efficacy of neither BBIVP-CorV vaccine nor the other authorized vaccines by WHO for emergency use has been identified yet among pwMS. This issue is of crucial value given the documented hindering effect of some DMTs – namely sphingosine 1-phosphate receptor (S1PR) modulators and anti-CD20 therapies – on proper immunization against COVID-19 (Achiron Anat et al. 2021; Achiron A., Mandel M., et al. 2021; Buttari et al. 2021; Chilimuri et al. 2021; Etemadifar Masoud; et al. 2021; Etemadifar M. et al. 2021; Gallo et al. 2021; Sormani et al. 2021). Accordingly, the effect of DMTs on practical vaccination efficacy in pwMS requires further investigation.

The reasons for vaccination hesitancy among the present study’s cohort of unvaccinated pwMS clarifies the role of misinformation in preventing pwMS from getting vaccinated. Providing evidence-based consultations and reasoning by their neurologists is an encouraged strategy to encounter misinformation among the pwMS. Setting mandates is the next possible option, although it may not be required if the pwMS are provided adequate evidence-based information.

In conclusion, the BBIVP-CorV COVID-19 vaccine seems relatively safe to administer in pwMS as it does not seem to evoke severe neurological symptoms or MS relapses. Like other COVID-19 vaccines, its efficacy among pwMS remains to be investigated in more controlled studies.

## Limitations

Several limitations apply to this work because of its remote nature: I) considering the limited framework of the study, we could not review the participants’ medical records to perform self-controlled analyses, II) the outcome measurements were performed in a self-reporting manner, which might not have been accurate, and III) the study lacked precise clinical and paraclinical evaluations.

## Data Availability

All anonymized data produced in the present study are available upon reasonable request from any qualified investigator to the authors.

## Disclosures

### Ethics

This study was approved by the ethics committee of Isfahan University of Medical Sciences. Considering the remote nature of the study, written consent could not be obtained from the participants, although they were completely informed about the aims of this study and all conveyed consent for anonymized publication of their information, both in the online surveys and on the telephone calls. No deanonymized data was stored by the investigators in any way, to ensure the privacy protection of the participants.

## Data Availability Statement

The anonymized data will be shared with any qualified investigator upon reasonable request.

